# Recommendations from long-term care reports, commissions, and inquiries in Canada

**DOI:** 10.1101/2020.11.17.20233114

**Authors:** Eric KC Wong, Trina Thorne, Carole Estabrooks, Sharon E Straus

**Affiliations:** Knowledge Translation Program, Li Ka Shing Knowledge Institute, St. Michael’s Hospital, 209 Victoria Street, Toronto, Ontario, M5B 1W8, Canada; Institute for Health Policy Management and Evaluation, Dalla Lana School of Public Health, University of Toronto, Ontario, Canada; Faculty of Nursing, University of Alberta, Edmonton, Alberta, T6G 1C9, Canada; Translating Research in Elder Care (TREC) program, University of Alberta, Edmonton, Alberta, T6G 1C9, Canada

**Author notes:** **Corresponding author:** Sharon Straus. **Competing interests:** The authors have no competing interests to declare.

## Abstract

**Background:** Multiple long-term care (LTC) reports over the last 30 years issued similar recommendations for improvement across Canadian LTC homes. Our primary objective was to identify the most common recommendations made over the past 10 years. Our secondary objective was to estimate the total cost of studying LTC issues repeatedly over the past 30 years.

**Methods:** The qualitative and cost analyses were conducted in Canada from July to October 2020. Using a list of reports, inquiries and commission from The Royal Society of Canada Working Group on Long-Term Care, we coded recurrent recommendations in LTC reports. We contacted the sponsoring organizations for a cost estimate, including direct and indirect costs. All costs were adjusted to 2020 Canadian dollar values.

**Results:** Of the 80 Canadian LTC reports spanning the years of 1998 to 2020, twenty-four (30%) were based on a national level and 56 (70%) were focused on provinces or municipalities. Report length ranged from 4 to 1491 pages and the median number of contributors was 14 (interquartile range, IQR, 5–26) per report. The most common recommendation was to increase funding to LTC to improve staffing, direct care and capacity (67% of reports). A median of 8 (IQR 3.25– 18) recommendations were made per report. The total cost for all 80 reports was estimated to be $23,626,442.78.

**Interpretation:** Problems in Canadian LTC homes and their solutions have been known for decades. Despite this, governments and non-governmental agencies continue to produce more reports at a monetary and societal cost to Canadians.

## Introduction

The COVID-19 pandemic led to a high proportion of deaths in Canadian long-term care homes (LTCH) compared with those of other developed countries. The proportion of deaths from COVID-19 in LTCH in Canada was 81% compared with a mean of 42% in other Organisation for Economic Cooperation and Development (OECD) countries [1]. This statistic is surprising since Canada is considered to have a relatively low number of COVID-19 deaths overall [2]. There were also significant differences between provinces that were attributed to pandemic preparedness, integration of services (LTCH, public health and hospitals), funding and resources, daily care hours for residents, comprehensiveness of inspections and differences in ownership model (profit versus non-profit) [3].

Residential LTCHs provide living accommodations for people who require full-time supervised care [4]. The founding of long-term care (LTC) in Canada was based in the Elizabethan Poor Law of 1601 that grouped older adults with other disadvantaged populations in housing that was run by various organizations and not centrally regulated [5]. LTC is not situated within the Canada Health Act [4], which resulted in different care models and resource allocation across organizations and homes. This variation has been associated with significantly different outcomes during the current pandemic [6].

During the early months of COVID-19, the media reported a shortage of direct care providers and personal protective equipment (PPE) in Canadian LTCHs, which led to residents suffering from a lack of basic personal care and delays in identifying medical problems [7]. Reduced staffing levels and wage compression in Canada’s LTC sector compelled individuals to work at more than one facility to make a living wage. An Ontario study comparing for-profit and non-profit facilities found lower staffing levels in for-profit LTCHs, which was associated with worse COVID-19 outcomes [8], although this may not be the case elsewhere in the country [9]. Working at multiple nursing homes contributed to the spread of COVID-19 infection [6], and although restricting employment to one facility reduced the number of outbreaks, it also exacerbated pre-existing work force shortages [10]. Furthermore, lower staffing levels and direct care hours are associated with increased rates of infection and hospital admission among residents [11], with this same trend observed in LTCHs with COVID-19 outbreaks [3,6,12].

In April, the Royal Society of Canada Working Group on Long-Term Care was tasked with reviewing the current state of LTC in the face of COVID-19 [5]. Their report found 103 LTC reports, commissions, and inquiries, 80 of which were unique reports based in Canada. The review found recurrent themes of longstanding deficiencies in the LTC sector that contributed to the magnitude of the COVID-19 crisis in LTCHs. Despite the wealth of evidence, little change has been implemented and in COVID’s wave 2, LTCHs are again being devastated by outbreaks [16]. LTCHs continue to account for the majority of outbreaks and deaths in Canada; as of Nov 7 there were 2,026 COVID-19 outbreaks in LTCHs [13]. The Canadian Red Cross has been called in to support care in 7 Ottawa LTCHs [14]. Since June, LTCHs across Canada have seen a 19% increase in the number of deaths [2]. The opportunity to support residents and care providers between wave 1 and wave 2 has passed. Sadly, the consequences of the lack of action will be disproportionately carried by the most vulnerable residents and the lowest paid, most marginalized members of the LTC workforce, personal support workers (PSW), who provide 90% of the direct care [15].

The primary **objective** of this study was to examine the recurring recommendations over the past 10 years. Our secondary objective was to calculate the total costs of generating all of the LTC reports, commissions, and inquiries in Canada over the last 30 years. We aimed to put the cost of repeatedly studying the same problems into context of the current pandemic.

## Methods

Qualitative and quantitative analyses were based on Canadian LTC reports and commissions identified from The Royal Society of Canada Working Group on Long-Term Care [5], which was done using a literature search and environmental scan over the last 30 years. Our analysis took place from July to October 2020. Content analysis was used to determine the presence of certain words, themes, or concepts within reports published in the past 10 years. Two reviewers independently reviewed 5 reports, and a consensus coding scheme was created. For the cost analysis, we excluded follow-up reports and included cost estimates in the original larger report.

### Estimation of report cost

We contacted the author or sponsoring organizations of the 80 Canadian LTC reports to inquire about the estimated cost of producing each report. We requested both direct and indirect costs. Direct costs included consultancy fees, salaries, compensation for expert witnesses, graphics, layout, printing, and dissemination. Indirect costs included time donated from authors who volunteered their time to produce the report. When an estimated budget was not available, we searched online for global budget reports from the sponsoring organization. Total annual expenses for research, advocacy, or reports were divided by the number of reports published that year by the organization to generate an estimated cost. We also searched for media reports about costs of commissions or coroner’s inquests. If no costs were available, the estimate was based on length, depth of research, inclusion of external experts/witnesses, and the sponsoring organization of similar LTC reports. Costs were in Canadian dollars and adjusted to 2020 values according to the Bank of Canada Inflation Calculator [16].

### Data abstraction and analysis

Report characteristics were extracted, including title, sponsoring organization, publication year, geographic region, scope of report, number of contributors, number of pages, and duration of the project. The primary outcome was recurrent recommendations. Secondary outcomes included contributors, total costs of producing the reports and median page count.

### Ethics approval

No ethics approval was required for this analysis.

## Results

The list of Canadian LTC reports from the Royal Society commission spans the years 1998 to 2020 (n=80). There was an increase in the number of reports over time, with 10 reports in the first half of 2020 (Figure 1). Most of the reports were focused at a provincial level (n=55, 68.8%), 24 reports were based on a national level (30.0%). We found one municipal report and no reports from the territories. Ontario (n=31, 55.0%) produced the majority of the provincial reports, followed by British Columbia (n=11, 19.6%) (Table 1). More reports were funded by provincial governments (n=26) compared with the federal government (n=9). Non-profit organizations (e.g. Canadian Association for Long Term Care, Canadian Institute of Health Information) and professional unions (e.g. Canadian Federation of Nurses Unions, Canadian Union of Public Employees) or associations (e.g. Registered Nurses’ Association of Ontario, Canadian Medical Association) authored the remaining 45 reports. No reports were funded by the private sector. Most of the reports focused solely on LTC (68.8%), but some reports focused on continuing care, older adults, or the health care system as well. The median report length was 40 pages (interquartile range, IQR, 21–84), with 16 reports (20.0%) over 100 pages. The median number of contributors, including authors, witnesses, and consultants, was 14 (IQR 5–26).

**Table 1:**
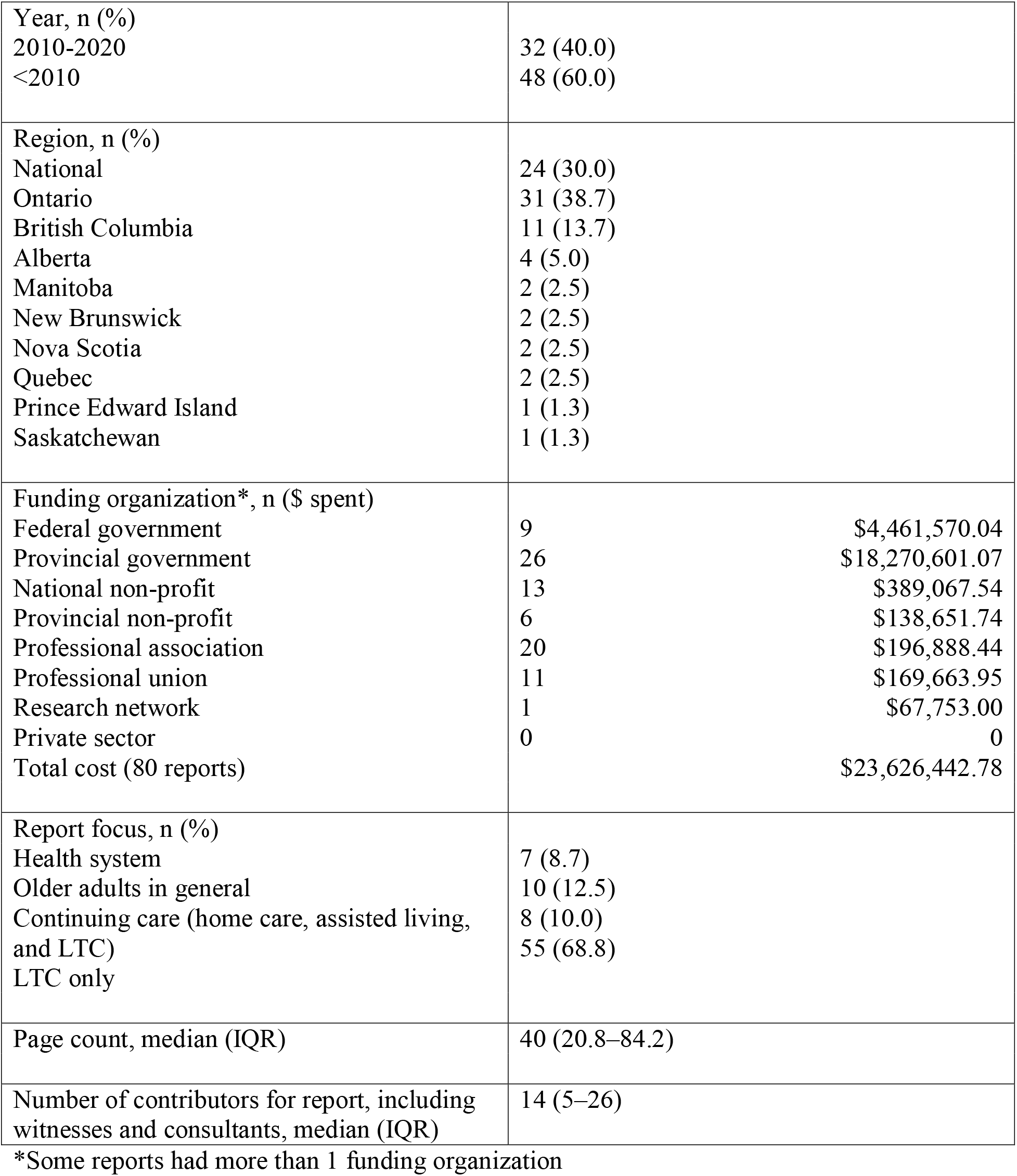
Report characteristics for 80 LTC reports in Canada from 1998-2020. LTC = long-term care; IQR = interquartile range

**Figure 1:**
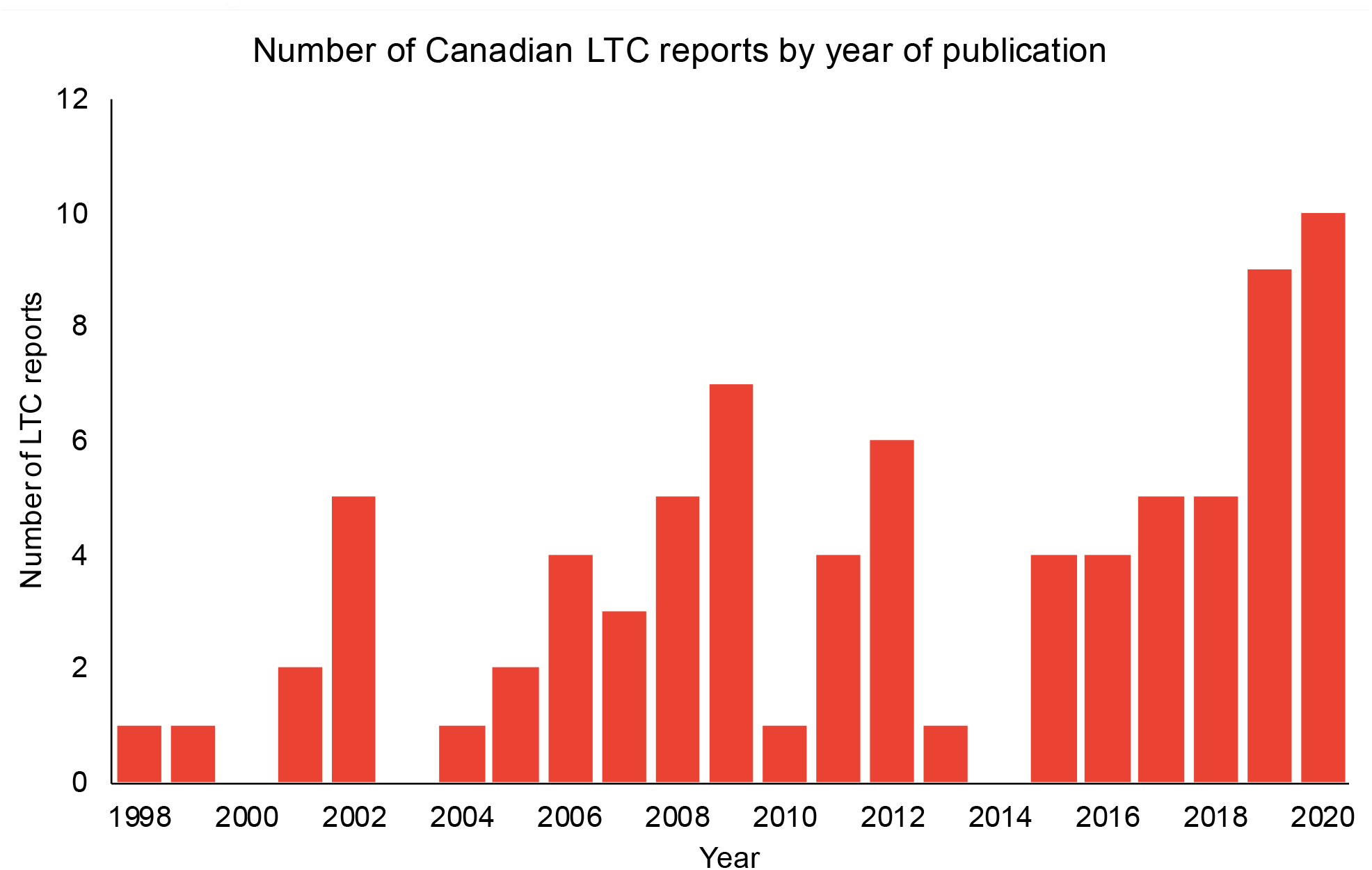
The number of Canadian reports with LTC recommendations over the span of 1998-2020. LTC = long-term care.

### Common recommendations in reports

Reviewing the reports from the last 10 years (n=48), we identified a median of 8 (IQR 3.25–18) recommendations per report. Numerous recommendations were repeated in these reports (Table 2). Overall, the most frequent recommendations were: (i) to increase or redistribute funding to improve staffing, direct care, and capacity (66.7%), (ii) to standardize, regulate and audit LTC quality of care (58.3%), and (iii) to standardize, regulate or reform education and training for LTC staff (52%). Improving staff education and training, increasing behavioural supports, and modernizing infection control measures were universally recommended in reports by governments, non-profits, professional association and unions.

**Table 2:**
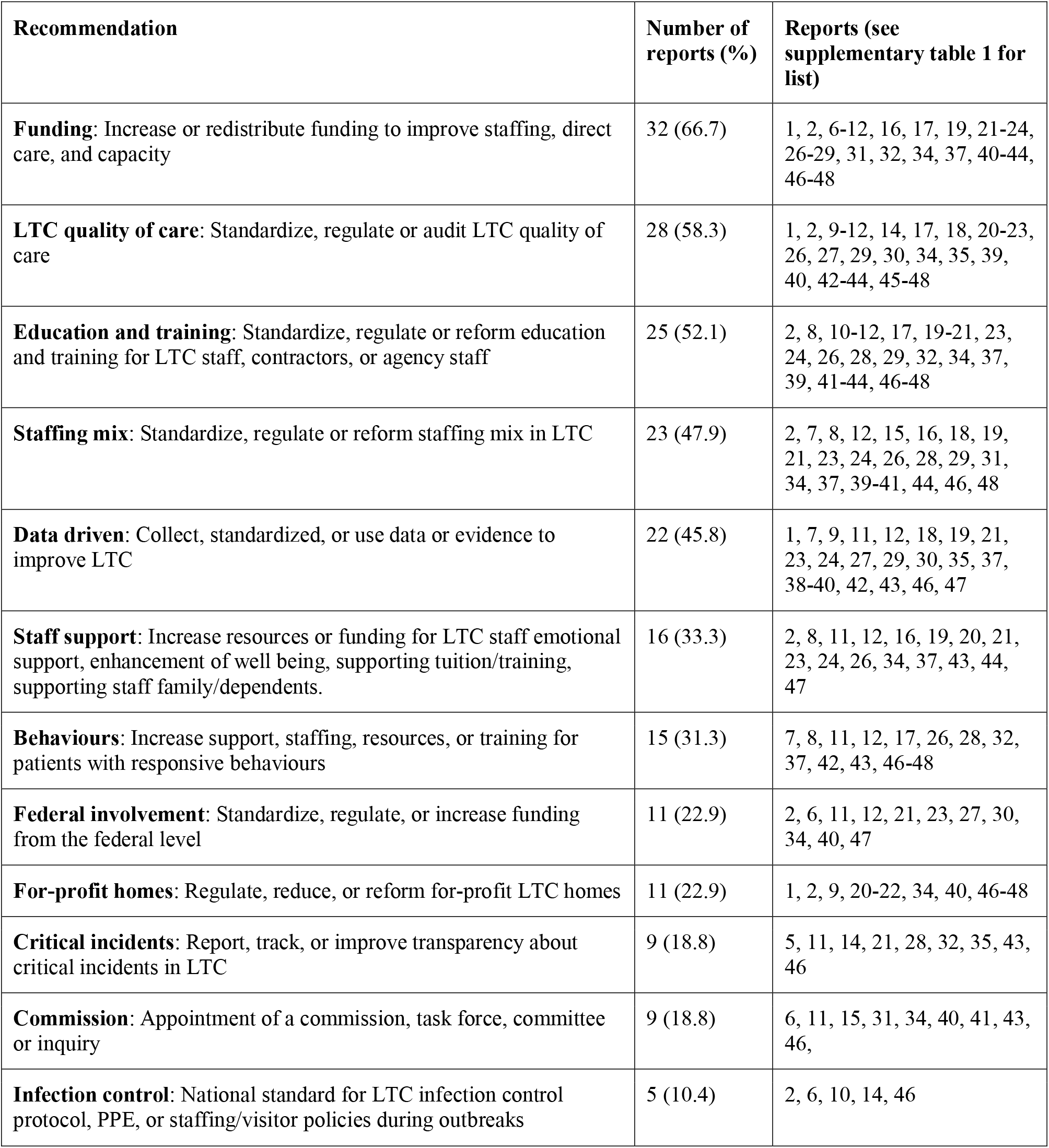
Recurrent recommendations in LTC reports in the years 2010-2020 (n=48). LTC = long-term care.

Government-authored reports (n=35) focused on using improving data collection (n=8, 22.9%), improving education and training of staff (n=8, 22.9%) and standardizing LTC quality of care (n=8, 22.9%). Professional union reports (n=11) focused on regulating for-profit LTCHs (n=4, 36.4%) and standardizing LTC quality of care (n=4, 36.4%). The most common recommendation made by non-profits (n=19) was to improve training and education for staff (n=9, 47.4%). Professional associations reports (n=20) most commonly recommended standardizing staffing mix (n=10, 50.0%) and increasing funding for direct care (n=10, 50.0%). The least common recommendation made by professional associations was for improved transparency, reporting and tracking of critical incidents (n=1, 5.0%).

Critical incidents received the most attention from governments (n=6, 17.1%), followed by non-profits (n=2, 10.5%) and a professional association (n=1, 5.0%). Recommendations regarding quality of care or data utilization were more likely to be made by government (n=18, 51.4%) or professional associations or unions (n=20, 64.5%) than by non-profits (n=3, 15.8%). Staff wellness support was most recommended by non-profits (n=7, 36.8%), followed by professional associations or unions (n=6, 19.4%), then government (n=3, 8.6%). The appointment of commissions or inquiries was mostly recommended by professional associations or unions (n=7, 22.6%) then by government (n=2, 5.7%) and no commissions were recommended by non-profits.

Although recommendations to improve resident quality of life were mentioned in 12 reports, they mostly overlapped with other recommendations such as increasing direct care, optimizing staff mix, increasing quality of life data collection, and improving quality of care. Only one report discussed specific recommendations to improve quality of life, such as increasing decision-making capacity (e.g. choice over when to bath), improving privacy with single rooms, and prioritizing relational care over medical tasks and interventions [17]. The full list of reports in Supplementary table 1.

### Cost of reports

Nearly half of the reports (45%) had cost estimates by the sponsoring organization or by publicly available budget or media reports. The total cost for all 80 reports was estimated to be $23,626,442.78 (Table 1). The median cost per report was $15,203.48 (IQR $10,000– $53,147.81). Details of each report’s cost estimate are shown in Supplementary table 2. The lowest cost was estimated at $500 for each of the Canadian Army Joint Task Force reports, which accounts for the administrative costs of writing the reports even if the services were provided on military order, but the cost of military deployment was not included [7]. The highest cost was $9,046,255.51 for the Public Inquiry into the Safety and Security of Residents in the Long-Term Care Homes System (The Wettlaufer Report) in 2019, which involved 79 contributors, witnesses, and experts [18].

## Discussion

This analysis highlights the efforts of multiple organizations, both governmental and non-governmental, to address the longstanding challenges and quality issues in Canada’s LTCHs.

The Royal Society Briefing Report concluded that the unresolved issues in Canada’s nursing homes have been known for decades. According to our analysis, there are numerous sound recommendations backed by evidence, yet little action has followed. This inaction has contributed to the devastating loss of life during the COVID-19 pandemic and to lower quality of life in LTCHs. Our analysis further shows the substantial cost of studying the problem repeatedly over the years.

Duplicate investigation of known findings reduces value and increases waste [19]. From the LTC reports, issues regarding understaffing, undertraining, and the negative impact of for-profit LTC homes were repeatedly mentioned [5]. Policy change often requires persistence [20], but the cost of advocating for change should be viewed from a societal context (financial and moral). Commissions and inquiries into LTC issues, like those happening or slated to begin around the country [21–23], can solidify our determination for policy change, but do not replace the need for policy implementation – action.

Several provinces have increased wages and provided full-time employment with more appropriate compensation and benefits to stabilize the LTC workforce [24]. The Ontario government went further to commit 4.00 hours per day of direct care for each LTC resident by 2024 [25], bringing the total care hours above the national average (3.30 hours) [26]. However, increasing direct care hours, while essential, is only one of the many recommendations from existing reports. Identifying the right staff mix and care team composition, providing proper education and training, and supporting staff wellness are also critical to developing a long-term workforce that has sufficient resilience to confront future crises [27]. Furthermore, we should focus more attention on resident quality of life, which should be the ultimate goal of our efforts.

Although the total cost of $23 million in generating LTC reports may seem insignificant compared to a government budget, it represents a lost opportunity to continually improve Canadian LTCHs. Studying the same problems repeatedly means Canadian experts are confined to fixing critical deficiencies in LTC instead of innovating new care models.

There are several strengths of this study. Two reviewers extracted key recommendations from each LTC report in the last 10 years and created a consensus coding scheme. We systematically tracked down the cost of each report by contacting the sponsoring organization or consulting their global budgets. The total costs, including time donated of experts authoring these reports, were accounted for. For reports that did not have available cost data, we estimated the total cost by using reports with known costs with similar length and depth.

The main limitation of this study was the lack of true cost estimates for half of the LTC reports. Some organizations lacked transparency about costing, and others lacked detailed accounting of spending. Staff turnover and record keeping practices were barriers to accessing data, particularly for the reports produced over 10 years ago. For government agencies, we often had to call multiple departments and speak with numerous representatives and noted considerable variation in the level of disclosure. For the two military reports during COVID-19, we likely underestimated the cost since they were generated as part of military duty. However, there were still considerable costs incurred by the public by having the military deployed to those LTCHs [28]. We also grouped reports on health system improvement and care of older adults in general in this analysis because LTC is intricately tied to the health system at large. Leaders in senior care such as Denmark also design LTC policy as in integral part of their health system [29].

## Conclusion

Over the last two decades, Canadian governments and non-government organizations have repeatedly investigated longstanding LTC issues and have largely drawn the same conclusions. Had the recurring recommendations been implemented, we would not only have improved working conditions, quality of care and quality of life, but would also have prevented unnecessary deaths due to COVID-19. Instead of continuing to investigate LTC issues, we should focus our resources on implementing the recommendations in the identified reports.

**Tables 1, 2 and Figure 1: separate file**

**Supplementary table 1: separate file**

**Supplementary table 2: separate file**

## Data Availability

Data not available for sharing.

