## Supplementary Table 1 for "Recommendations from long-term care reports, commissions, and inquiries in Canada"

**Supplementary table 1**: Canadian reports with LTC recommendations with report number linked with table 2 in the main text. LTC = long-term care; BC = British Columbia.

| **Report #** | **Year** | **Report title** |
| --- | --- | --- |
| 1* | 2020 | Long-Term Care in Ottawa: We Need Change Now, A Statement of Concerns |
| 2 | 2020 | Re-imagining Long-term Residential Care in the COVID-19 Crisis |
| 3 | 2020 | 4th Canadian Division Joint Task Force (Central). OP LASER – JTFC Observations in Long Term Care Facilities in Ontario |
| 4 | 2020 | OBSERVATIONS SUR LES CENTRES D’HÉBERGEMENT DE SOINS LONGUES DURÉES DE MONTRÉAL |
| 5 | 2020 | Protecting Human Rights During and After COVID-19: Challenges to the human rights of older people in Canada |
| 6 | 2020 | 2020 Vision: Improving long-term care for people in Canada |
| 7 | 2020 | Enhancing Community Care for Ontarians (ECCO) 3.0 |
| 8 | 2020 | Caring in Crisis: Ontario’s Long-Term Care PSW Shortage, Report & recommendations from the front lines across Ontario |
| 9 | 2020 | A Billion Reasons to Care: A Funding Review of Contracted Long-Term Care in B.C. |
| 10 | 2020 | COVID-19 Infection Prevention and Control Practices in Long-Term Care, Residential and Retirement Homes |
| 11 | 2019 | The Public Inquiry into the Safety and Security of Residents in the Long-Term Care Homes System (The Wettlaufer Report) |
| 12 | 2019 | Enabling the Future Provision of Long-Term Care in Canada |
| 13 | 2019 | The Future Co$t of Long-Term Care in Canada |
| 14 | 2019 | Food and Nutrition in Long-Term-Care Homes |
| 15 | 2019 | A better approach to long-term care in Ontario |
| 16 | 2019 | This is Long-Term Care 2019: The impact of dementia, New evidence about quality of care, The need for more staff |
| 17 | 2019 | Planning, Access, Levels of Care and Violence in Ontario’s Long-Term Care |
| 18 | 2019 | Filling the Gap: Determining Appropriate Staffing & Care Levels for Quality in Long Term Care |
| 19 | 2019 | Minister’s Expert Advisory Panel on Long-Term Care: Recommendations |
| 20 | 2018 | Negotiating Tensions in Long-Term Residential Care |
| 21 | 2018 | Ensuring Quality Care For All Seniors |
| 22 | 2018 | Crumbling Away: Saskatchewan Long-Term Residential Care Policy and Its Consequences |
| 23 | 2018 | Accelerating our innovation potential: Actions to advance innovation in Ontario’s long-term care system. |
| 24 | 2018 | The Future of Long-Term Care is Now: Addressing nursing care needs in Manitoba’s Personal Care Homes |
| 25* | 2017 | Sizing Up the Challenge: Meeting the Demand for Long-Term Care in Canada |
| 26 | 2017 | Exercising Choice in Long-Term Residential Care |
| 27 | 2017 | Caring for Canada’s Seniors: Recommendations for meeting the needs of an aging population |
| 28 | 2017 | Aging with Confidence Ontario Action Plan for Seniors |
| 29 | 2017 | Residential Care Staffing Review |
| 30 | 2016 | The State of Seniors health Care in Canada |
| 31 | 2016 | Mind the Safety Gap in Health System Transformation: Reclaiming the Role of the RN |
| 32 | 2016 | Resident to Resident Aggression in B.C. Care Homes |
| 33 | 2016 | Private seniors’ residences: more than just rental businesses |
| 34 | 2015 | Before It’s Too Late: A National Plan for Safe Seniors’ Care |
| 35 | 2015 | Long-Term-Care Home Quality Inspection Program Standing Committee on Public Accounts |
| 36 | 2015 | Strengthening Seniors Care delivery in BC |
| 37 | 2015 | Broken Homes: Nurses speak out on the state of long-term care in Nova Scotia and chat a course for a sustainable future |
| 38 | 2013 | When a Nursing Home Is a Home: How Do Canadian Nursing Homes Measure Up on Quality? |
| 39 | 2012 | WHY NOT NOW? A Bold, Five-Year Strategy for Innovating Ontario’s System of Care for Older Adults |
| 40* | 2012 | From Bad to Worse: Residential Elder Care in Alberta |
| 41 | 2012 | Submission to the Government of Ontario’s Seniors Care Strategy |
| 42 | 2012 | Living Longer, Living Well Report. Submitted to the Minister of Health and Long-Term Care and the Minister Responsible for Seniors on recommendations to Inform a Seniors Strategy for Ontario |
| 43 | 2012 | Long-Term Care Task Force on Resident Care and Safety. An Action Plan to Address Abuse and Neglect in Long-Term Care Homes |
| 44* | 2011 | Element of an Effective Innovation Strategy for long Term Care in Ontario |
| 45 | 2011 | Response to the Ontario Seniors’ Secretariat on: Phase Two of the Initial Draft Regulations under the Retirement Homes Act, 2010 |
| 46 | 2011 | Response to the Ontario Seniors’ Secretariat on: Initial Draft Regulations under the Retirement Homes Act |
| 47 | 2011 | Long-Term Care in Canada: Status Quo No Option |
| 48 | 2010 | Position Statement: Strengthening Client Centered Care in Long-Term Care; 2010 |

* Reports that have 2 funding sources

**Source:** Estabrooks CA, Straus S, Flood, CM, Keefe J, Armstrong P, Donner G, Boscart V, Ducharme F, Silvius J, Wolfson M. Restoring trust: COVID-19 and the future of long-term care. Royal Society of Canada. 2020.
