## Supplementary Table 2 for "Recommendations from long-term care reports, commissions, and inquiries in Canada"

**Supplementary table 2**: All Canadian health reports with LTC recommendations and estimated cost for each report. The estimated actual cost includes direct and indirect costs, such as time donated by authors and monetary inflation to 2020 values. Justification for the estimated cost is included in the rightmost column. Global budget indicates reverse calculation of costs from global research budget for the organization in the year of publication, divided by the number of publications that year. LTC = long-term care; BC = British Columbia.

| Title | Year | Region | Name of primary funding organization | Funding Source | Primary Focus | Authors | Page count | Budget Accessibility | Total cost of report | Estimated actual cost with inflation | Key factors in estimating cost |
| --- | --- | --- | --- | --- | --- | --- | --- | --- | --- | --- | --- |
| Annual Report: Long-term Care Community Services Activity | 1998 | Ontario | Auditor General of Ontario | Provincial Government | Residential long-term care |  | 22 | Budget accessed online | $416,405.00 | $625,063.09 | Global budget |
| Nursing Task Force, Ontario. Good Nursing, Good Health: An Investment for the 21st Century | 1999 | Ontario | Ministry of Health & Long-term Care | Provincial Government | Health System | 17 | 25 | Contacted- nonresponse |  | $20,000.00 | Detailed policy report |
| A Framework for Reform, Report of the Premier’s Advisory Council on Health (The Mazankowski Report) | 2001 | Alberta | Government of Alberta (Premier’s Advisory Council on Health for Alberta) | Provincial Government | Health System | 18 | 75 | Budget accessed online | $2,000,000.00 | $2,788,617.89 | Media identified budget; inflated to 2020 value |
| Report of a study to review level of service and responses to need in a sample of long term care facilities and selected comparators | 2001 | Ontario | Ontario Long-Term Care Association | Provincial Non Profit, Professional Association | Residential long-term care |  | 100 | Contacted-response- budget request denied |  | $50,000.00 | Detailed report; well researched and analyzed |
| Commission on the Future of Health Care in Canada. Building on Values: The Future of Health Care in Canada. | 2002 | Canada | Committee of the Privy Council | Federal Government | Health System | 35 | 474 | Details found through other sources (e.g. media) | $15,000,000.00 | $4,013,611.94 | Media reported; inflated to 2020 value; 19.6% of the report was on aging and LTC |
| The Health of Canadians – The Federal Role Final Report Volume Six: Recommendations for Reform (Kirby report) | 2002 | Canada | Standing Senate Committee on Social Affairs, Science and Technology | Federal Government | Health System | 40 | 392 | Budget estimated from future reports. No annual report from that year. | $20,000.00 | $50,000.00 | Extensive report; numerous witnesses; time donated |
| Ownership Matters: Lessons Learned from Long-Term Care Facilities | 2002 | BC | Hospital Employees’ Union (B.C.) | Professional Union | Residential long-term care |  | 26 | Contacted- nonresponse |  | $10,000.00 | Moderate length; well researched |
| Jumping on the Alberta Bandwagon: Does B.C. need this kind of Assisted Living? | 2002 | BC | Hospital Employees’ Union (B.C.) | Professional Union | Continuing Care (LTC, Assisted living & home care) | 1 | 9 | Contacted-response- budget details provided | $10,000.00 | $13,651.74 | Author's estimate |
| Eldercare: on the auction block | 2002 | Alberta | The Alberta Chapter Consumers’ Association of Canada | Provincial Non Profit | Residential long-term care | 1 | 38 | Contacted-response- budget details provided | $10,000.00 | $13,651.74 | Author's estimate; inflated |
| Commitment to Care: A Plan for Long-Term Care in Ontario | 2004 | Ontario | Ministry of Health & Long-term Care | Provincial Government | Residential long-term care | 49 | 60 | Contacted- nonresponse |  | $30,000.00 | Detailed report; stakeholder consultation |
| CONTINUING CARE RENEWAL OR RETREAT? BC Residential and Home Health Care Restructuring 2001–2004 | 2005 | BC | Canadian Centre for Policy Alternatives | National Non Profit | Continuing Care (LTC, Assisted living & home care) | 14 | 54 | Not contacted; add on study |  | $25,000.00 | Moderate report size, multiple researchers |
| Report of the Auditor General on Seniors Care and Programs | 2005 | Alberta | Auditor General of Alberta | Provincial funds-Auditor General | Continuing Care (LTC, Assisted living & home care) |  | 97 | Contacted-response- budget details provided | Figure not available (10,868 staff hours + $215,000 contractor fees) | $541,040.00 | From hourly estimate x $30/hr for analyst min wage plus contractor fees |
| NACA demands improvement to Canada's long term care institutions | 2006 | Canada | National Advisor y Council on Aging | National Non Profit | Older Adults in general |  | 10 | Organization dissolved. |  | $2,500.00 | Brief advocacy statement with citation of existing research |
| Long-term Care In Manitoba | 2006 | Manitoba | Manitoba Nurses Union | Professional Union | Residential long-term care |  | 32 | Contacted- nonresponse |  | $10,000.00 | Internal report; some research; moderate length |
| Residential care quality: A review of the literature on nurse and personal care staffing and quality of care | 2006 | BC | Nursing Directorate British Columbia Ministry of Health | Provincial Government | Residential long-term care | 12 | 85 | Contacted- nonresponse |  | $50,000.00 | Detailed report; well researched; broad consultation |
| Report on the inquest into the deaths of Ezzeldine El Roubi and Pedro Lopez | 2006 | Ontario | Office of the Chief Coroner, Ontario | Provincial Government | Residential long-term care | 26 | 107 | Contacted-response- budget details provided |  | $200,000.00 | Assume cost of inquest plus witnesses; detailed report; |
| First Interim Report, Embracing the Challenge of Aging | 2007 | Canada | Special Senate Committee on Aging | Federal Government | Older Adults in general | 39 | 104 | Budget accessed online | $98,991.00 | $121,263.97 | Global budget; averaging report cost; inflated |
| Staffing and Care Standards for Long-Term Care Homes | 2007 | Ontario | Registered Nurses' Association of Ontario | Professional Association | Residential long-term care |  | 12 | Contacted-response- budget request denied |  | $5,000.00 | Brief policy statement |
| Dignity Denied: Long-Term Care and Canada’s Elderly | 2007 | Canada | National Union of Public and General Employees (NUPGE) | Professional Union | Residential long-term care |  | 52 | Contacted- nonresponse |  | $7,000.00 | Moderate details; broad scope |
| Interim Report, Issues and Options for an Aging Population | 2008 | Canada | Special Senate Committee on Aging | Federal Government | Older Adults in general | 16 | 34 | Budget accessed online | $98,991.00 | $117,284.67 | Global budget; averaging report cost; inflated |
| Policy Brief #4, HHRP issues: A series of policy options the long-term care environment: Improving outcomes through staffing decisions | 2008 | Canada | Canadian Nurses Association | Professional Association | Residential long-term care |  | 4 | Contacted, budget not available due to age of report | $8,504.48 | $10,100.00 | Estimated from another similar report from same organization |
| Final Summary Report: Trends, Projections and Recommended Approaches to Delivery of Long-term Care in the Province of Prince Edward Island 2007-2017 | 2008 | PEI | PEI Department of Health and Wellness | Provincial Government | Residential long-term care | 2 | 21 | Contacted- nonresponse |  | $10,000.00 | Policy statement; moderate length; no consultation stated |
| Be independent. Longer. New Brunswick’s Long-Term Care Strategy | 2008 | New Brunswick | The Government of New Brunswick | Provincial Government | Residential long-term care |  | 36 | Contacted- nonresponse |  | $10,000.00 | Policy statement; moderate length; no consultation stated |
| People Caring for People: Impacting the Quality of Life and Care of Residents of Long-Term Care Homes | 2008 | Ontario | Saint Elizabeth Health Care | Provincial Non Profit | Residential long-term care | 99 | 84 | Contacted-response- budget details provided | $0.00 | $50,000.00 | Detailed research; broad consultation; time donated by key author |
| Special Senate Committee on Aging: Final Report, Canada’s Aging Population: Seizing the opportunity | 2009 | Canada | Senate Committee on Aging | Federal Government | Older Adults in general | 16 | 247 | Budget accessed online | $98,991.00 | $118,409.46 | Global budget; averaging report cost |
| New Directions for Facility-Based Long Term Care | 2009 | Canada | Canadian Healthcare Association | National Non Profit | Residential long-term care | 5 | 172 | Contacted-response- budget details provided | $12,862.00 | $15,385.06 | Global budget; internal authors; no consultations |
| Response to the Minister of Health and Long-Term Care on: Part 2 of the Draft Regulation under the Long-Term Care Homes Act, 2007 | 2009 | Ontario | Registered Nurses’ Association of Ontario | Professional Association | Residential long-term care |  | 18 | Contacted-response- budget request denied |  | $5,000.00 | Brief policy statement |
| Response to the Minister of Health and Long-term Care on: Initial Draft Regulation under the Long-Term Care Homes Act, 2007 | 2009 | Ontario | Registered Nurses’ Association of Ontario | Professional Association | Residential long-term care |  | 19 | Contacted-response- budget request denied |  | $5,000.00 | Brief policy statement |
| Infection Prevention and Control at Long-term-care Homes | 2009 | Ontario | Auditor General of Ontario | Provincial Government | Residential long-term care |  | 27 | Budget accessed online | $685,175.00 | $819,581.60 | Global budget; inflated |
| The Best of Care: Getting it Right for Seniors in British Columbia: Part 1 | 2009 | BC | The B.C. Ombudsman | Provincial Government | Continuing Care (LTC, Assisted living & home care) | 12 | 70 | Contacted- nonresponse |  | $20,000.00 | Broad consultation; review of cases; detailed report |
| Being There for Seniors: Our Progress in Long-Term Care, New Brunswick's long-term care strategy | 2009 | New Brunswick | The Government of New Brunswick | Provincial Government | Residential long-term care |  | 33 | Contacted- nonresponse |  | $10,000.00 | Policy statement; moderate length; no consultation stated |
| Position Statement: Strengthening Client Centered Care in Long-Term Care; 2010 | 2010 | Ontario | Registered Nurses’ Association of Ontario | Professional Association | Residential long-term care |  | 12 | Contacted-response- budget request denied |  | $5,000.00 | Brief policy statement |
| Element of an Effective Innovation Strategy for long Term Care in Ontario | 2011 | Ontario | The Conference Board of Canada | National Non Profit, Professional Association | Residential long-term care | 11 | 94 | Contacted- nonresponse |  | $50,000.00 | Detailed report; extensive research |
| Response to the Ontario Seniors’ Secretariat on: Initial Draft Regulations under the Retirement Homes Act | 2011 | Ontario | Registered Nurses' Association of Ontario | Professional Association | Continuing Care (LTC, Assisted living & home care) |  | 15 | Contacted-response- budget request denied |  | $5,000.00 | Brief advocacy statement |
| Response to the Ontario Seniors’ Secretariat on: Phase Two of the Initial Draft Regulations under the Retirement Homes Act, 2010 | 2011 | Ontario | Registered Nurses' Association of Ontario | Professional Association | Continuing Care (LTC, Assisted living & home care) |  | 8 | Contacted-response- budget request denied |  | $5,000.00 | Brief advocacy statement |
| Long-Term Care in Canada: Status Quo No Option | 2011 | Canada | Canadian Federation of nurses Unions | Professional Union | Residential long-term care | 19 | 112 | Contacted-response- budget request denied |  | $7,000.00 | Time donate; interviews of various stakeholders; mainly descriptive; long report |
| Submission to the Government of Ontario’s Seniors Care Strategy | 2012 | Ontario | Registered Nurses’ Association of Ontario | Professional Association | Older Adults in general |  | 27 | Contacted-response- budget request denied |  | $10,000.00 | Moderate length report; research but some based on prior reports |
| From Bad to Worse: Residential Elder Care in Alberta | 2012 | Alberta | Parkland Institute (Faculty of Arts, University of Alberta) | Professional Union, Research Network | Residential long-term care |  | 72 | Contacted-response- budget details provided | $60,000.00 | $67,753.09 | Author estimate; inflated |
| WHY NOT NOW? A Bold, Five-Year Strategy for Innovating Ontario’s System of Care for Older Adults | 2012 | Ontario | Ontario Trillium Foundation, Government of Ontario | Provincial Government | Residential long-term care | 22 | 120 | No budget found, due to lack of organization ownership. |  | $100,000.00 | Detailed report; high profile author panel; project duration |
| Long-Term Care Task Force on Resident Care and Safety. An Action Plan to Address Abuse and Neglect in Long-Term Care Homes | 2012 | Ontario | Long-Term Care Task Force on Resident Care and Safety | Provincial Government | Residential long-term care | 50 | 110 | Contacted-response- budget request denied |  | $1,000,000.00 | Detailed report; extensive consultation and research; not at the scale of the commission reports; chair says most is time donated |
| Living Longer, Living Well Report. Submitted to the Minister of Health and Long-Term Care and the Minister Responsible for Seniors on recommendations to Inform a Seniors Strategy for Ontario | 2012 | Ontario | Ministry of Seniors of Ontario | Provincial Government | Older Adults in general | 422 | 233 | Contacted-response- budget request denied |  | $100,000.00 | Very detailed report; extensive consultation and research; time donated |
| The Best of Care: Getting it Right for Seniors in British Columbia: Part 1 | 2012 | BC | The B.C. Ombudsman | Provincial Government | Residential long-term care | 26 | 254 | Contacted- nonresponse |  | $50,000.00 | Broad consultation; review of cases; detailed report; updated in 2019 |
| When a Nursing Home Is a Home: How Do Canadian Nursing Homes Measure Up on Quality? | 2013 | Canada | Canadian Institute of Health Information (CIHI) | Federal Government | Residential long-term care | 9 | 34 | Contacted-response- budget request denied |  | $15,000.00 | Small team data analysis; medium size erport |
| Strengthening Seniors Care delivery in BC | 2015 | BC | BC Care Providers Association | Professional Association | Continuing Care (LTC, Assisted living & home care) | 1 | 59 | Contacted-response- budget details provided | $15,000.00 | $16,166.54 | Author provided costs of similar reports as reference |
| Before It’s Too Late: A National Plan for Safe Seniors’ Care. | 2015 | Canada | Canadian Federation of Nurses Unions | Professional Union | Continuing Care (LTC, Assisted living & home care) | 3 | 58 | Contacted-response- budget request denied |  | $10,000.00 | Time donated; no external consultation; moderate length |
| Broken Homes: Nurses speak out on the state of long-term care in Nova Scotia and chat a course for a sustainable future | 2015 | Nova Scotia | Nova Scotia Nurses Union | Professional Union | Residential long-term care | 8 | 76 | Contacted-response- budget details provided | $0.00 | $20,000.00 | Broad consultation; time donated |
| Long-Term-Care Home Quality Inspection Program Standing Committee on Public Accounts | 2015 | Ontario | Auditor General of Ontario | Provincial Government | Residential long-term care |  | 37 | Budget accessed online | $805,093.00 | $867,704.32 | Global budget; inflated |
| The State of Seniors health Care in Canada | 2016 | Canada | Canadian Medical Association | Professional Association | Older Adults in general | 6 | 20 | Contacted-response- budget details provided | $0.00 | $10,000.00 | Time donated; detailed research across multiple provinces |
| Mind the Safety Gap in Health System Transformation: Reclaiming the Role of the RN | 2016 | Ontario | Registered Nurses' Association of Ontario | Professional Association | Health System |  | 76 | Contacted-response- budget request denied |  | $20,000.00 | Detailed advocacy statement; moderate research/analysis |
| Resident to Resident Aggression in B.C. Care Homes | 2016 | BC | Office of the Seniors Advocate British Columbia | Provincial Government | Residential long-term care |  | 30 | Contacted-response- budget details provided | $380,000.00 | $404,468.58 | Global budget; inflated |
| Private seniors’ residences: more than just rental businesses | 2016 | Quebec | The Quebec ombudsman | Provincial Government | Residential long-term care |  | 2 | Contacted- nonresponse |  | $5,000.00 | Based on average ombudsman case spending in other provinces; no budget from Quebec |
| Exercising Choice in Long-Term Residential Care | 2017 | Canada | Canadian Centre for Policy Alternatives | National Non Profit | Residential long-term care | 50 | 128 | Contacted-response- budget details provided | $14,006.00 | $50,000.00 | Time donated by experts and authors (LTC visits, 2M grant) |
| Sizing Up the Challenge: Meeting the Demand for Long-Term Care in Canada | 2017 | Canada | The Conference Board of Canada | National Non Profit, Professional Association | Residential long-term care | 4 | 48 | Contacted- nonresponse |  | $40,000.00 | Detailed report, economic modelling and projections; extensive research and analysis |
| Caring for Canada’s Seniors: Recommendations for meeting the needs of an aging population | 2017 | Canada | Canadian Association for Long Term Care | Professional Association | Residential long-term care |  | 6 | Contacted- nonresponse |  | $2,500.00 | Brief advocacy statement with citation of existing research |
| Residential Care Staffing Review | 2017 | BC | British Columbia Ministry of Health | Provincial Government | Residential long-term care |  | 44 | Contacted- nonresponse |  | $10,000.00 | Report of moderate length; no consultations |
| Aging with Confidence Ontario Action Plan for Seniors | 2017 | Ontario | Ministry of Seniors of Ontario | Provincial Government | Older Adults in general |  | 39 | Contacted-response- budget request denied |  | $10,000.00 | In-house general review; brief report |
| Negotiating Tensions in Long-Term Residential Care | 2018 | Canada | Canadian Centre for Policy Alternatives | National Non Profit | Residential long-term care | 50 | 136 | Contacted-response- budget details provided | $15,754.00 | $50,000.00 | Time donated by experts and authors (LTC visits, 2M grant) |
| Ensuring Quality Care For All Seniors | 2018 | Canada | Canadian Health Coalition | National Non Profit | Older Adults in general |  | 25 | Contacted- nonresponse |  | $5,000.00 | Brief report; time donated |
| Accelerating our innovation potential: Actions to advance innovation in Ontario’s long-term care system. | 2018 | Ontario | Ontario Long-Term Care Association | Professional Association | Residential long-term care | 23 | 35 | Contacted-response- budget request denied |  | $35,000.00 | Report of moderate length; high profile consultants |
| Crumbling Away: Saskatchewan Long-Term Residential Care Policy and Its Consequences | 2018 | Saskatchewan | CUPE Health Care Workers | Professional Union | Residential long-term care | 4 | 40 | Contacted-response- budget details provided | $12,000.00 | $12,259.12 | Organization declared |
| The Future of Long-Term Care is Now: Addressing nursing care needs in Manitoba’s Personal Care Homes | 2018 | Manitoba | Manitoba Nurses Union | Professional Union | Residential long-term care |  | 40 | Contacted- nonresponse |  | $12,000.00 | Internal report; well researched; moderate length |
| Enabling the Future Provision of Long-Term Care in Canada | 2019 | Canada | National Institute on Ageing | National Non Profit | Residential long-term care | 18 | 161 | Contacted-response- budget details provided | $62,500.00 | $62,591.24 | Author estimated, including time donated |
| The Future Co$t of Long-Term Care in Canada | 2019 | Canada | National Institute on Ageing | National Non Profit | Residential long-term care | 14 | 45 | Contacted-response- budget details provided | $62,500.00 | $62,591.24 | Author estimated, including time donated |
| Filling the Gap: Determining Appropriate Staffing & Care Levels for Quality in Long Term Care | 2019 | BC | BC Care Providers Association | Professional Association | Residential long-term care | 1 | 51 | Contacted-response- budget details provided | $15,000.00 | $15,021.90 | Author provided costs of similar reports as reference |
| This is Long-Term Care 2019: The impact of dementia, New evidence about quality of care, The need for more staff | 2019 | Ontario | Ontario Long-Term Care Association | Professional Association | Residential long-term care |  | 16 | Contacted-response- budget request denied |  | $7,000.00 | Brief report of available statistics |
| A better approach to long-term care in Ontario | 2019 | Ontario | Registered Nurses' Association of Ontario | Professional Association | Residential long-term care |  | 4 | Contacted-response- budget request denied |  | $1,000.00 | Brief statement based on prior research |
| The Public Inquiry into the Safety and Security of Residents in the Long-Term Care Homes System (The Wettlaufer Report) | 2019 | Ontario | Attorney General of Ontario | Provincial Government | Residential long-term care | 79 | 1491 | Contacted-response- budget details provided | $9,000,000.00 | $9,000,000.00 | Based on government internal estimate |
| Food and Nutrition in Long-Term-Care Homes | 2019 | Ontario | Auditor General of Ontario | Provincial Government | Residential long-term care |  | 47 | Budget accessed online | $1,056,328.00 | $1,057,870.08 | Global budget |
| Minister’s Expert Advisory Panel on Long-Term Care: Recommendations | 2019 | Nova Scotia | Queen’s Printer for Ontario | Provincial Government | Residential long-term care | 3 | 26 | Contacted- nonresponse |  | $15,000.00 | Detailed research and some consultation |
| Planning, Access, Levels of Care and Violence in Ontario’s Long-Term Care | 2019 | Ontario | Ontario Health Coalition | Provincial Non Profit | Residential long-term care | 10 | 44 | Contacted-response- budget details provided | $0.00 | $15,000.00 | Time donated; well researched |
| OBSERVATIONS SUR LES CENTRES D’HÉBERGEMENT DE SOINS LONGUES DURÉES DE MONTRÉAL | 2020 | Quebec | 2nd Canadian Division Joint Task Force (East) | Federal Government | Residential long-term care |  | 60 | Contacted-response- budget details provided | $0.00 | $500.00 | Reporting on observations; indirect administrative cost of report writing |
| 4th Canadian Division Joint Task Force (Central). OP LASER – JTFC Observations in Long Term Care Facilities in Ontario | 2020 | Ontario | 4th Canadian Division Joint Task Force | Federal Government | Residential long-term care | 1 | 15 | Contacted-response- budget details provided | $0.00 | $500.00 | Reporting on observations; indirect administrative cost of report writing |
| Long-Term Care in Ottawa: We Need Change Now, A Statement of Concerns | 2020 | Ontario | The Council of Aging of Ottawa | Federal Government, Provincial Government, Provincial Non Profit | Residential long-term care | 6 | 20 | Contacted-response- budget details provided | $25,000.00 | $25,000.00 | Author estimated, including time donated |
| Re-imagining Long-term Residential Care in the COVID-19 Crisis | 2020 | Canada | Canadian Centre for Policy Alternatives | National Non Profit | Residential long-term care | 5 | 16 | Contacted-response- budget details provided | $0.00 | $1,000.00 | Brief report; time donated by experts |
| COVID-19 Infection Prevention and Control Practices in Long-Term Care, Residential and Retirement Homes | 2020 | Canada | Accreditation Canada | National Non Profit | Health System |  | 12 | Contacted- nonresponse |  | $10,000.00 | Internal report; average complexity; brief report |
| Protecting Human Rights During and After COVID-19: Challenges to the human rights of older people in Canada | 2020 | Canada | INTERNATIONAL LONGEVITY CENTRE CANADA | National Non Profit | Older Adults in general | 5 | 12 | Contacted-response- budget details provided | $15,000.00 | $15,000.00 | Time donated; estimated by authors |
| 2020 Vision: Improving long-term care for people in Canada | 2020 | Canada | Canadian Nurses Association | Professional Association | Residential long-term care |  | 13 | Contacted-response- budget details provided | $10,100.00 | $10,100.00 | From authors |
| Enhancing Community Care for Ontarians (ECCO) 3.0 | 2020 | Ontario | Registered Nurses' Association of Ontario | Professional Association | Health System | 22 | 92 | Contacted-response- budget request denied |  | $30,000.00 | Detailed report on health system level change; some research; mostly advocacy |
| A Billion Reasons To Care: A Funding Review of Contracted Long-Term Care in B.C. | 2020 | BC | Office of the Seniors Advocate British Columbia | Provincial Government | Residential long-term care |  | 51 | Contacted-response- budget details provided | $480,000.00 | $480,000.00 | Global budget |
| Caring in Crisis: Ontario’s Long-Term Care PSW Shortage, Report & recommendations from the front lines across Ontario | 2020 | Ontario | Ontario Health Coalition | Provincial Non Profit | Residential long-term care |  | 34 | Contacted-response- budget details provided | $0.00 | $10,000.00 | Time donated; moderate length report |
